# Transcriptional survey of peripheral blood links lower oxygen saturation during sleep with reduced expressions of CD1D and RAB20 that is reversed by CPAP therapy

**DOI:** 10.1101/19001727

**Authors:** Tamar Sofer, Ruitong Li, Roby Joehanes, Honghuang Lin, Adam C. Gower, Heming Wang, Nuzulul Kurniansyah, Brian E. Cade, Jiwon Lee, Stephanie Williams, Reena Mehra, Sanjay R. Patel, Stuart F. Quan, Yongmei Liu, Jerome I. Rotter, Stephen S. Rich, Avrum Spira, Daniel Levy, Sina A. Gharib, Susan Redline, Daniel J. Gottlieb

## Abstract

Sleep Disordered Breathing (SDB) is associated with a wide range of physiological changes due, in part, to the influence of hypoxemia during sleep. We studied gene expression in peripheral blood mononuclear cells in association with three measures of SDB: Apnea Hypopnea Index (AHI); average oxyhemoglobin saturation (avgO2) during sleep; and minimum oxyhemoglobin saturation (minO_2_) during sleep. We performed discovery association analysis in two community-based studies, the Framingham Offspring Study (FOS; N=571) and the Multi-Ethnic Study of Atherosclerosis (MESA; N = 580). An association with false discovery rate (FDR) *q <* 0.05 in one study was considered “replicated” if a *p* < 0.05 was observed in the other study. Those genes that replicated across MESA and FOS, or with FDR *q <* 0.05 in meta-analysis, were used for analysis of gene expression in the blood of 15 participants from the Heart Biomarkers In Apnea Treatment (HeartBEAT) trial. HeartBEAT participants had moderate or severe obstructive sleep apnea (OSA) and were studied pre- and post-treatment (three months) with continuous positive airway pressure (CPAP). We also performed Gene Set Enrichment Analysis (GSEA) on all traits and cohort analyses. Twenty-two genes were associated with SDB traits in both MESA and FOS. Of these, lower expression of *CD1D* and *RAB20* was associated with lower avgO2 in MESA and FOS. CPAP treatment increased the expression of these genes in HeartBEAT participants. Immunity and inflammation pathways were up-regulated in subjects with lower avgO2; *i.e*., in those with a more severe SDB phenotype (MESA), whereas immuno-inflammatory processes were down-regulated in response to CPAP treatment (HeartBEAT).

**One Sentence Summary:** We studied the association of gene expression in blood with obstructive sleep apnea traits, including oxygen saturation during sleep, and identified mechanisms that are reversed by treatment with Continuous Positive Airway Pressure.

## Introduction

Sleep Disordered Breathing (SDB) is characterized by abnormal respiratory patterns during sleep and is usually associated with impaired gas exchange. The most common type, Obstructive Sleep Apnea (OSA), is estimated to be prevalent in 17% of women and 34% of men, aged 30-70 years (1), and contributes significantly to the incidence and morbidity of cardio-metabolic disease, including hypertension, stroke, heart failure, coronary artery disease, atrial fibrillation, and diabetes (2-8). OSA is characterized by episodic complete (apneas) or partial (hypopneas) cessation of breathing during sleep, resulting in intermittent hypercapnic hypoxemia. Hypoxemia, particularly in the presence of hypercapnia, leads to both acute and persistent increases in sympathetic nervous system activity (9) that persists during wakefulness (10, 11) and contributes to elevated blood pressure. The arousal that typically occurs at the termination of apneas and hypopneas further increases sympathetic activity. Intermittent hypoxemia is also implicated in systemic inflammation and oxidative stress that further increases risk for cardio-metabolic disease (12-14). The molecular mechanisms underlying OSA-mediated physiological stress are not well established, but include genetic and gene regulatory factors.

Genome-wide gene expression analysis is a powerful tool for identifying molecular mechanisms underlying complex traits (15-17). Hypoxemia is known to influence gene expression, partly by affecting transcription factor activation, including both HIF-1α and NF-κB (18); therefore, the impact of OSA on gene expression, and particularly that of inflammatory genes, is anticipated. Although gene expression profiles may differ across tissues (19), intermittent hypoxemia has been shown to affect gene expression in peripheral blood mononuclear cells of healthy volunteers (20). Currently, studies of gene expression in OSA (or SDB) are limited. Five studies ranging in size from 18 to 48 individuals have evaluated the impact of OSA on gene expression in peripheral blood monocytes or total leukocytes, either cross-sectionally in comparison to those without OSA (20-22), or pre- and post-treatment with continuous positive airway pressure (CPAP) (23, 24), the most common therapy for OSA. These small studies had limited power to detect significant expression differences (or changes with treatment) in individual genes, although pathway analysis suggested possible involvement of inflammatory, cell cycle, and neoplastic pathways (23). In addition to their small sample size, some studies targeted a specific patient population (*e*.*g*., children) (22) or a limited set of genes (21), and generally focused on dichotomized (presence/absence) OSA traits. Larger sample sizes and analysis of quantitative OSA trait values are important to increase statistical power, enhance generalizability of the results, and provide insight into which aspects of OSA may be responsible for observed effects on gene expression. Complementary analyses that address causality are needed to overcome confounding that may be present in observational studies.

To address these gaps in knowledge, we conducted the largest analysis to date of genome-wide transcriptional associations with multiple traits that characterize different aspects of SDB: the apnea hypopnea index (AHI) as well as the average and the minimum oxyhemoglobin saturation during sleep (avgO2 and minO2, respectively). We analyzed this large, population-based peripheral blood transcriptomics data set in relation to SDB phenotypes using two complementary approaches with respect to: gene-focused analysis with cross validation, and pathway-based methods. For the gene-focused approach, we first performed a discovery association analysis of gene expression in two population-based cohorts: the Framingham Offspring Study (FOS, (25)) and the Multi-Ethnic Study of Atherosclerosis (MESA, (26)). Second, we further evaluated genes identified in the discovery study using fifteen individuals from the Heart Biomarkers in Apnea Treatment (HeartBEAT) clinical trial (27). These subjects had moderate to severe OSA and were adherent to CPAP for three months. In the pathway-based approach, we performed Gene Set Enrichment Analysis (GSEA) in each of these cohorts to determine those processes that are associated with distinct SDB traits and, in the case of HeartBEAT, we assessed the effect of OSA therapy in altering canonical biological pathways.

## Materials and Methods

The central study design is illustrated in Fig. 1. We start with a single-gene association discovery step, in which we studied the association of gene-expression with SDB traits in two discovery cohorts: the Framingham Offspring Study (FOS) and the Multi-Ethnic Study of Atherosclerosis (MESA). First, we performed association analysis in each of FOS and MESA separately, and tested for replication of gene expression associations in the second study (cross-replication), following the standard in genetic analysis for replication as a gold standard for limiting false discoveries. Then, to increase power, we meta-analyzed estimated associations across FOS and MESA. Finally, we took genes that either cross-replicated between FOS and MESA or were detected in their meta-analysis and studied where the expression of these genes responds to CPAP treatment in the HeartBEAT clinical study.

**Fig. 1.**
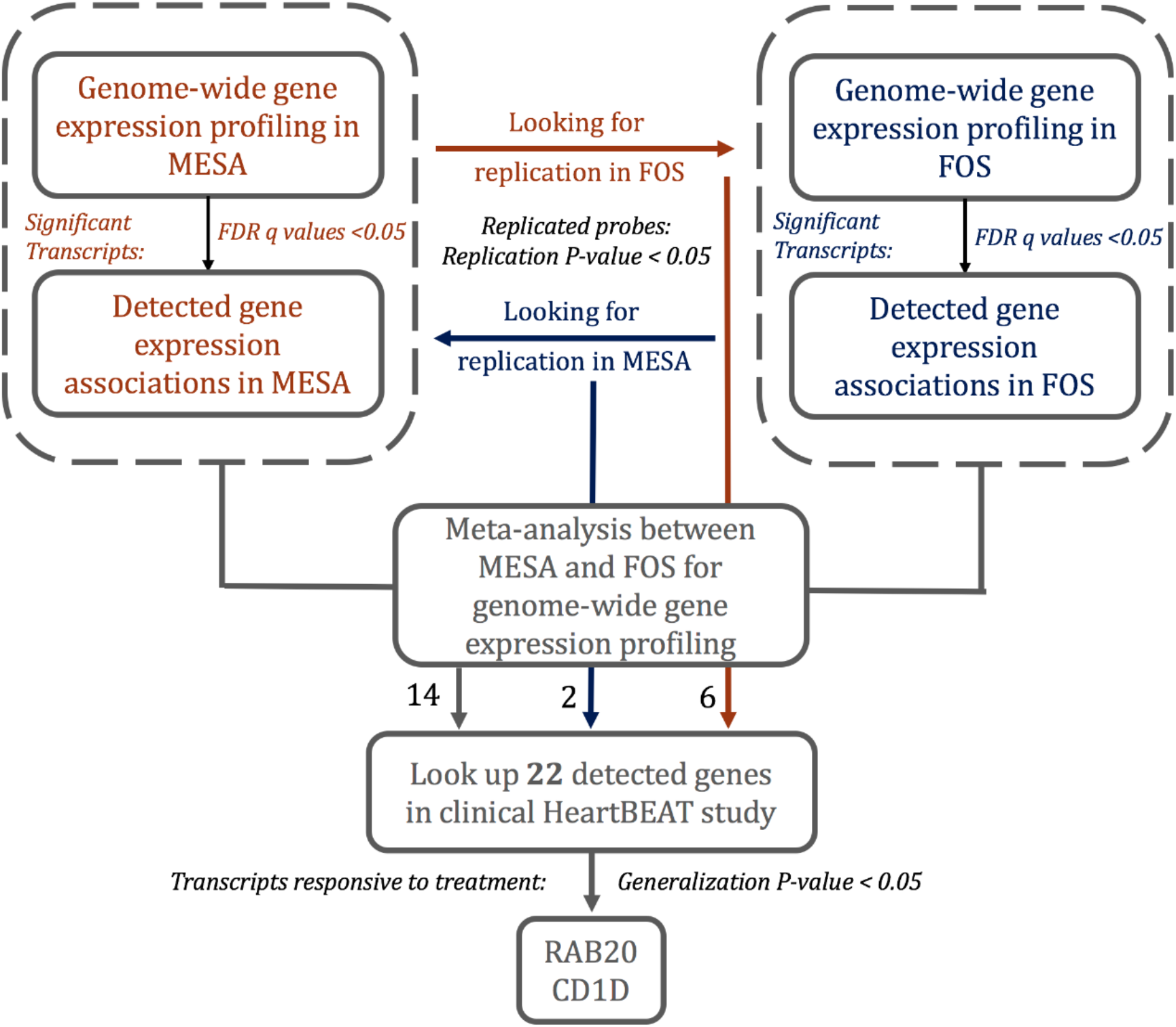
Gene-based analysis workflow. This figure displays the cross-replication and meta-analysis approach for discovery of gene transcripts associated with OSA traits, and follow-up association analysis of response to CPAP treatment in HeartBEAT.

### The Multi-Ethnic Study of Atherosclerosis (MESA)

MESA is a longitudinal cohort study, established in 2000, that prospectively collected risk factors for development of cardiovascular disease among participants in six field centers across the United States (Baltimore City and Baltimore County, MD; Chicago, IL; Forsyth County, NC; Los Angeles County, CA; Northern Manhattan and the Bronx, NY; and St. Paul, MN). The cohort has been studied every few years. For the present analysis, we consider N = 619 individuals who participated in a sleep study (28, 29) and had available gene expression data. Blood for gene expression was drawn during the Exam 5 core examination (2010-2012) and sleep studies were conducted in an ancillary exam (2010-2013). Blood collection and monocyte sample purification were performed sequentially using sodium heparin-containing Vacutainer CPT_TM_ cell separation tubes (Becton Dickinson, Rutherford, NJ, USA) and anti-CD14-coated magnetic beads, using an AutoMACs automated magnetic separation unit (Miltenyi Biotec, Bergisch Gladbach, Germany), as detailed before (30). Sleep data were collected using standardized full in-home level-2 polysomnography (Compumedics Somte Systems, Abbotsville, Australia, AU0), as described before (29). After removal of 39 observations due to missing sleep data, we included 580 participants in the analysis: 196 African-Americans (AA), 259 European-Americans (EA) and 125 Hispanic-Europeans (HA).

### Gene expression data collection and processing in MESA

Gene expression profiling in MESA was described in Liu et al (30). Briefly, peripheral blood samples of 1,264 randomly selected MESA participants from four MESA field centers (Baltimore, MD; Forsyth County, NC; New York, NY; and St Paul, MN) were obtained during the fifth examination, and mononuclear cells were isolated with consistent purity of > 90%. Blood was drawn in the morning following 8 hours fast. Extracted RNA that passed Quality Control (QC) testing was reversely transcribed, amplified and subjected to global expression microarrays. Global expression quantification was measured by Illumina HumanHT-12 v4 Expression BeadChip and Illumina Bead Array Reader. Samples were assigned to chips via stratified random sampling to prevent potential batch, chip, and position biases. Gene expression data underwent pre-processing, QC, and normalization, in steps including local background correction, elimination of probes not satisfying QC criteria, and normal-exponential convolution model. This was done using a series of Bioconductor packages (beadarray, limma etc) and Illumina software GenomeStudio. These steps resulted in gene expression data for 14,619 transcripts for further statistical analysis.

### Association analyses in MESA

We fit linear mixed models separately in each race/ethnic group to estimate the association between SDB exposures and gene expression markers, with a random effect for gene expression chip. Covariates (fixed effects) were study site, residual cell contamination (Neutrophils, Natural Killer cells, T cell, B cell), age, and sex. When the model did not converge due to null variance component, we fit a linear regression model with original fixed effects. The regression results from the three groups where then combined using inverse-variance fixed effects meta-analysis. In sensitivity analysis, we also adjusted for BMI.

### Framingham Offspring Study (FOS)

FOS, initiated in 1971, is a prospective epidemiologic study of 5,124 young adults, consisting of offspring of the original Framingham Heart Study cohort and their recruited spouses (31). The cohort has been examined every 4-8 years for standardized medical history, blood tests and 12-lead ECG (32, 33). Of FOS participants, 699 were enrolled in the Sleep Heart Health Study (SHHS), which was initiated with a first visit (1995 – 1998) that included in-home level-2 polysomnography (Compumedics P-Series, Abbotsville, Australia), followed by a second visit (2001 – 2003), in which a subset of 385 people underwent repeat polysomnography (34, 35). At the FOS eighth examination cycle (2005 - 2008), RNA was extracted from whole blood for gene expression profiling (36, 37). In this manuscript, we consider 517 individuals, all European-Americans, with both gene expression and polysomnography data. To minimize the time gap between gene expression and polysomnography measurements and maximize statistical power, the polysomnography measurements and corresponding phenotypes were taken from the second SHHS visit when available, and otherwise, from the first SHHS visit.

### Gene expression data collection and processing in FOS

Gene expression profiling and consecutive processing for FOS cohort have been described in detail by Joehanes et al and Lin et al (37, 38). Briefly, collected peripheral whole blood samples from each participant were used for RNA extraction. Total RNA was then amplified, labeled, and reversely transcribed into cDNA. The cDNA hybridization was conducted on a Human Exon 1.0 ST array. Pre-processing steps included log2 transformation, quantile normalization, and summarization with Robust Multi-array Average method. QC steps excluded exons with low signal to noise ratio and transcript clusters with missing matches in RefSeq transcript records. This resulted in 17,873 distinct transcripts available for analysis. Subsequent statistical analysis was adjusted for multiple batch effects and technical variables and estimated white cell counts as described in (37).

### Association analysis in FOS

We fit linear mixed models to estimate the association between SDB exposure and gene expression markers, with a random effect accounting for familial relatedness. Covariates (fixed effects) were estimated white blood cell counts (Neutrophils, Natural Killer cells, T cell, B cell), sex, age, and in sensitivity analysis, BMI. Because for FOS the expression measures were taken from blood 2-13 years after the sleep study, it is not clear how and if time varying covariates may confound the SDB-trait gene expression association, and we adjusted for age and BMI measures at the time of the sleep study.

### SDB phenotypes and covariates

In both discovery cohorts, we studied three quantitative measures of SDB, assessed by overnight PSG: 1) Apnea-Hypopnea indexes (AHI), defined as the number of apneas plus hypopneas per hour of sleep that were associated with at least 3% oxygen desaturation from the preceding baseline; 2) Average oxyhemoglobin saturation during sleep (avgO2); and 3) Minimum oxyhemoglobin saturation (minO2) during sleep. Covariates included sex, age, and BMI (for sensitivity analysis).

### Cross-replication analysis

Each of MESA and FOS studies where treated as both a “discovery” and a “replication” study. For each discovery cohort and each of the SDB exposures, we computed FDR-adjusted *p* values (FDR *q* values; (39)) across the expression transcripts. Then, we took all expression transcripts with *q* < 0.05 for replication in the other cohort. Replication is complicated by the fact that there are sometimes multiple transcripts per gene, and further, MESA and FOS used different platforms, so that gene transcripts did not necessarily match between the discovery and replication studies. Therefore, we used an approach termed “many to many” (40) in which a transcript of a specific gene in the discovery study was matched to all transcripts of the same gene in the replication study. After matching the list of discovery transcripts with FDR *q* < 0.05 in one cohort to a list of transcripts in the second cohort, a corresponding gene was replicated if one of the transcripts in the replication study had a one-sided *p* value < 0.05, requiring that the direction of association be the same as the transcript in the discovery study (41). Replicated genes were carried forward for further analysis in the CPAP treatment cohort.

### Meta-analysis of MESA and FHS

Using the same “many-to-many” approach described above, we matched all expression transcripts in MESA and FOS, and performed inverse-variance fixed-effects meta-analysis for overall exposure effect. We computed FDR-controlling *q*s. Genes with at least one transcript with FDR-value < 0.05 were carried forward for further analysis in the CPAP treatment cohort.

### Treatment response in a clinical cohort: the Heart Biomarker Evaluation in Apnea Treatment (HeartBEAT) study

The Heart Biomarker Evaluation in Apnea Treatment (HeartBEAT) study is a randomized, 4-site single-blind clinical trial that investigated the efficacy of OSA therapy in reducing cardiovascular disease risk for patients with moderate-severe OSA (ClinicalTrials.gov NCT01086800) (27). (27). Of HeartBEAT participants randomized to the CPAP treatment group, a subsample of 15 individuals who were highly adherent to CPAP therapy (defined by at least 4 hours of CPAP use per night over the 3-month intervention period) was selected for gene expression analysis. Venipuncture was performed in the morning following an overnight fast. Venous blood was collected in 8 mL heparinized Cell Prep Tubes containing Ficoll Hypaque (Becton Dickinson #362753) in order to separate peripheral blood mononuclear cells. The tubes were centrifuged fresh at room temperature for 15 minutes at 2000 G to isolate the buffy coat, which was pelleted, resuspended in Millipore S-002-10F freezing medium, and cryopreserved at −80C. Total RNA was extracted using Qiagen’s miRNeasy Mini kit following the manufacturer’s standard protocol. Gene expression profiling was performed using the Affymetrix Human Gene 1.0 ST array in samples collected before initiating CPAP and after three months of CPAP treatment. Raw CEL files were normalized to produce gene-level expression values using the implementation of the Robust Multiarray Average (RMA) in the affy Bioconductor R package (version 1.36.1) and an Entrez Gene-specific probeset mapping from the Molecular and Behavioral Neuroscience Institute (Brainarray) at the University of Michigan (version 16.0.0). Student’s paired *t* test was performed using the implementation in the multtest R package (version 2.14.0), and correction for multiple hypothesis testing was accomplished using the Benjamini-Hochberg false discovery rate (FDR). Analysis of the HeartBEAT microarray data was performed using the R environment for statistical computing (version 2.15.1).

### Testing of genes identified by MESA and FOS analyses

We used a statistical replication testing framework to study whether genes associated with SDB traits respond to OSA treatment using CPAP. We calculated the mean fold change with gene expression data collected post- and pre-CPAP therapy across the 15 HeartBEAT subjects. For any given gene that was carried forward from either the cross-replication or the meta-analysis of MESA and FOS (see Fig. 1), we computed the replication *p* value by applying a one-sided t-test on the log of the estimated fold expression change. Because the replication framework, requires that effects are consistent in their direction in the discovery and the replication stage, we defined the direction of association between gene expression marker and OSA as positive if the mean fold change was larger than 1 and negative otherwise. Thus, we computed one-sided *p* values so that a gene could be declared as replicated in HeartBEAT only if its expression was positively associated with more severe SDB in MESA and FOS and decreased following three months of CPAP treatment in HeartBEAT (or vice versa). Finally, a gene was determined as responding to treatment if its replication (one-sided) p value was < 0.05.

### Gene Set Enrichment Analysis

We performed gene set enrichment analysis (GSEA, (42)) for quantitative SDB traits in each cohort. For MESA and FOS, GSEA was applied separately for AHI, avgO2, and minO2 using the entire gene expression profiles rank ordered based on each gene’s test statistic for the corresponding SDB trait. For HeartBEAT, the transcriptome was rank ordered based on the paired *t* statistic computed with respect to CPAP therapy. When more than one transcript was available for a single gene (for MESA and FOS), a single transcript was selected at random. To maximize biological relevance, we leveraged only curated pathways (Molecular Signature Database “Canonical pathways” and “Hallmark” collections, gene symbol identifiers, version 6.0) for a total of 1379 gene sets, and performed permutation analysis (n = 1000 permutations). We used cutoff of FDR *q* < 0.05 to identify significantly enriched pathways. We used Enrichment Map (43), a plug-in application within the Cytoscape (v3.3.1) environment (44), to develop a network-based visualization of the GSEA results.

## Results

### Participant Characteristics

Demographic characteristics of the discovery studies FOS and MESA and the clinical intervention study HeartBEAT are provided in Table 1. Age and BMI were similar across cohorts. As expected, HeartBEAT study participants, who were recruited based on a minimum AHI of 15 events/hour, were somewhat heavier and had more severe OSA than the general population samples of FOS, and were also almost entirely male. FOS and HeartBEAT participants were almost all of European descent, while the MESA participants included many of African and Hispanic/Latino ancestry. Fig. 1 summarizes the analysis workflow.

**Table 1.** Characteristics of participants in the MESA, FOS, and HeartBEAT studies. For continuous variables, the table provides mean and standard deviation (in parentheses). For categorical variables, the table provides number of participants and percent of cohort (in parentheses). FOS characteristics are at the time of the sleep study. HeartBEAT characteristics are prior to CPAP treatment, except for CPAP adherence, which provides the average number of hours used by the participant per night over the three-months treatment period.

| Characteristics | Cohort |  |  |
| --- | --- | --- | --- |
|  | MESA | FOS | HeartBEAT |
| N | 619 | 517 | 15 |
| **Demographics** |  |  |  |
| Age (yr) | 68.7 (9.2) | 62.21 (9.0) | 65.85 (7.7) |

| **Characteristics** | **Cohort** |  |  |
| --- | --- | --- | --- |
|  | **MESA** | **FOS** | **HeartBEAT** |
| Male sex (%) | 329 (53.2) | 264 (51.1) | 14 (93.3) |
| Ethnicity (%) |  |  | - |
| African Americans (%) | 130 (21.0) | - | - |
| Hispanic Americans (%) | 203 (32.8) | - | - |
| European Americans (%) | 286 (46.2) | 517 (100.0) | 13 (86.7) |
| Mixed (%) | - | - | 2 (13.3) |
| BMI (kg/m<sup>2</sup>) | 29.81 (5.6) | 29.02 (5.3) | 31.45 (3.3) |
| **OSA Traits** |  |  |  |
| avgO2 (%) | 94.2 (1.8) | 94.3 (2.0) | 93.3 (1.7) |
| minO2 (%) | 83.0 (7.7) | 84.5 (6.2) | 77.7 (7.2) |
| AHI (events/ hr) | 19.6 (18.9) | 14.4 (15.1) | 36.9 (9.4) |
| CPAP adherence (hr/night) | -- | -- | 5.7 (1.9) |

### Gene-based identification of transcripts associated with SDB traits

To identify differentially expressed genes associated with SDB traits, we first performed a cross-replication analysis, in which we conducted discovery analysis in each of FOS and MESA, followed by replication analysis in the other cohort, approach aligned with the current standards in genetic studies considered vital to interpretation of statistically significant findings. We then meta-analyzed the information from FOS and MESA to increase power (45).

Table 2 lists eight genes that associated with the SDB traits in the cross-replication analysis. The expression of *RAB20* was positively associated with avgO2, while expression of *CDYL* was negatively associated with avgO2, in both FOS and MESA. Moreover, *RAB20* expression increased following CPAP treatment in HeartBEAT (*p* = 0.046). The expression of *TUBB6, INVS1ABP, MAPK1, VIM, STX2*, and *CRIP1* was associated with either minO2 or avgO2 in both MESA and FOS, but their expression was unchanged following CPAP treatment in HeartBEAT.

**Table 2.**
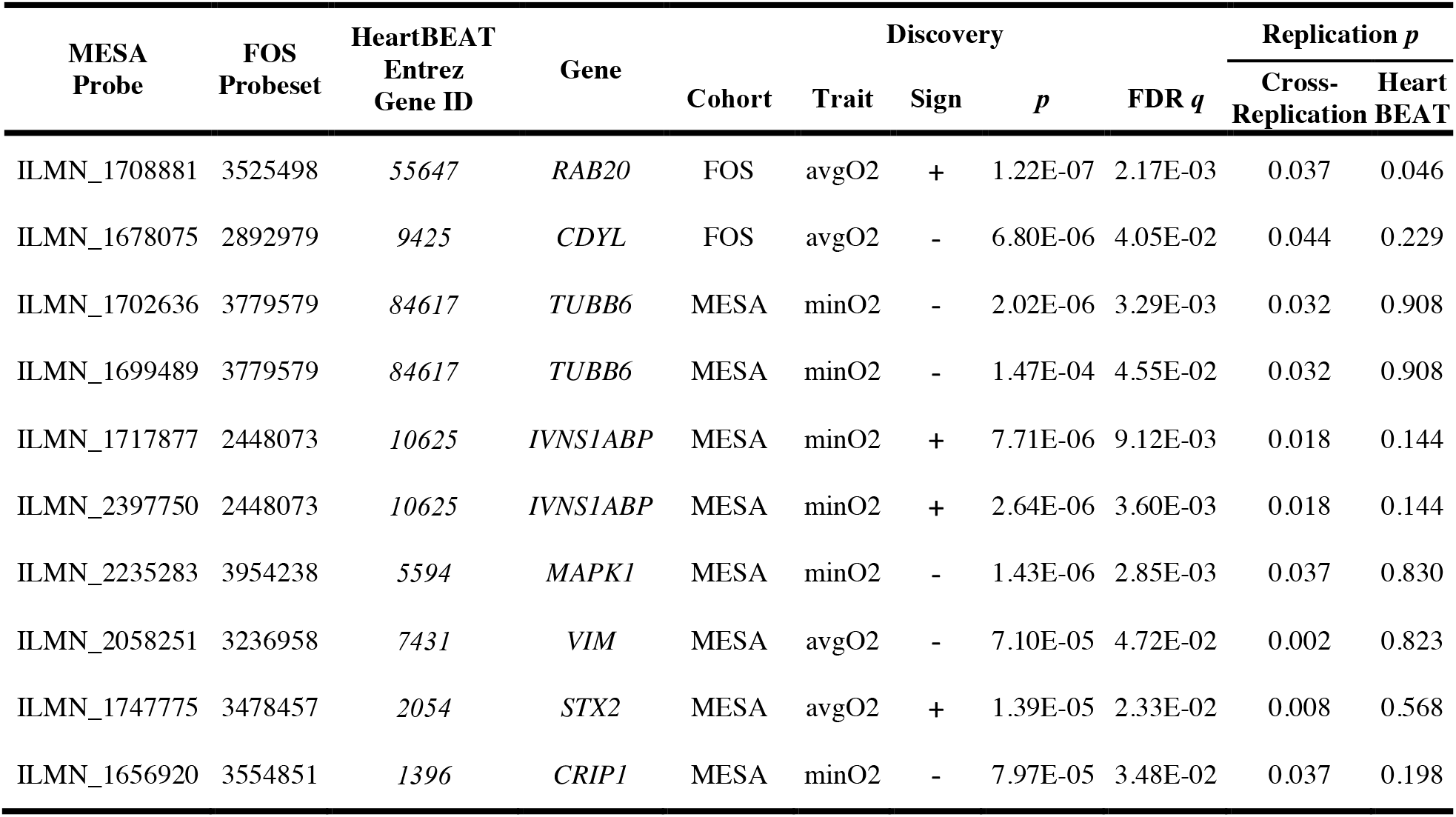
Genes identified in cross-replication analysis of MESA and FOS. Each row represents a gene transcript that replicated between FOS and MESA. Discovery cohort is the study in which the expression transcript was discovered (FDR *q <* 0.05). The identifiers corresponding to the transcript within each gene expression platform (MESA probe, FOS probeset, HeartBEAT Entrez Gene ID) are indicated. Discovery *p* is the original discovery *p* value, prior to multiple-testing adjustment. Discovery FDR-value is the FDR *q* in the discovery study, after FDR correction was applied to the discovery trait analysis. Replication *p* values are one-sided *p* values in the follow-up cohorts, with directions based on the direction of association in the discovery cohort. Cross-replication refers to FOS, when MESA is the discovery study, and to MESA, when FOS is the discovery study.

Table 3 lists 21 genes whose expressions was associated with either AHI, avgO2, or minO2 in the meta-analysis of FOS and MESA. Of these genes, 14 (67%) were significant only in the meta-analysis, including *CD1D*, whose expression was lower in association with more severe OSA in the discovery cohorts and significantly increased with CPAP in the HeartBEAT study (*p* = 0.002). Fig. 2 provides a clustered heatmap of the effect sizes (in standard deviations) of the identified transcripts that either cross-replicated between FOS and MESA, or were detected in the meta-analysis for each of the SDB traits.

**Table 3.**
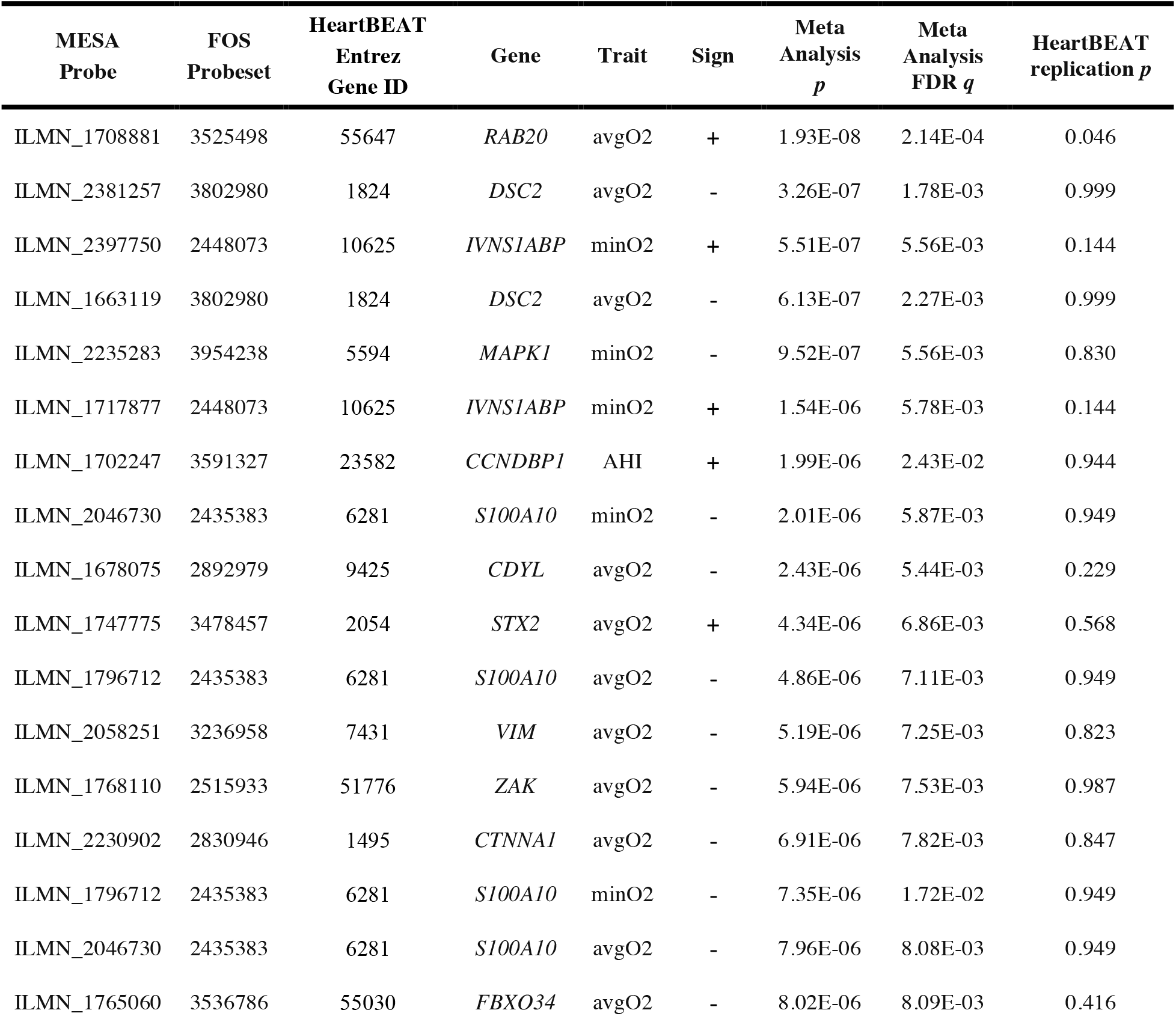
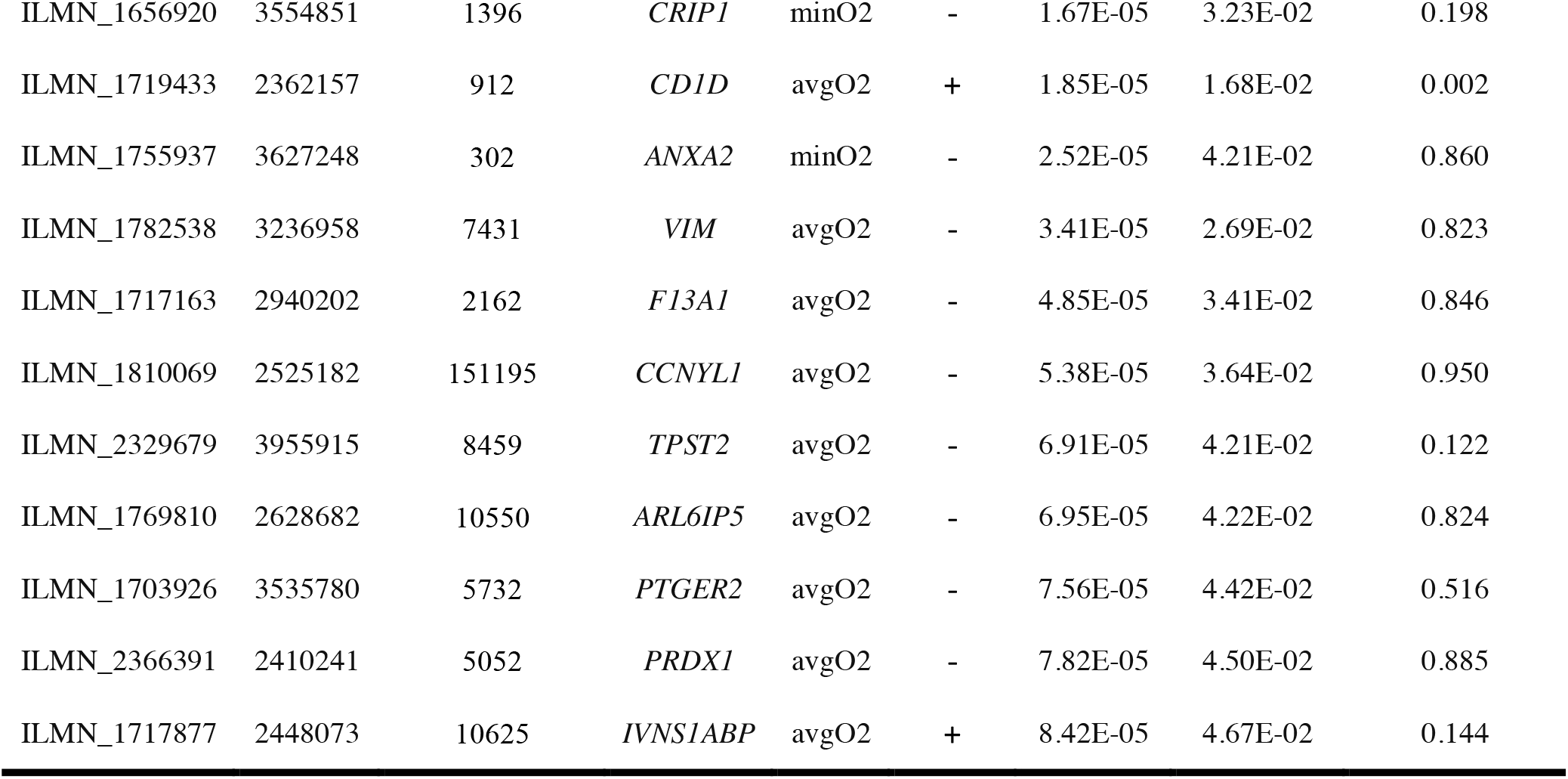
Genes identified in the meta-analysis of FOS and MESA. Each row represents a gene transcript that was identified (FDR *q <* 0.05) in the meta-analysis of FOS and MESA trait-specific results. The identifiers corresponding to the transcript within each gene expression platform (MESA probe, FOS probeset, HeartBEAT Entrez Gene ID) are indicated. Meta-analysis *p* values and FDR *q* values are the original and the FDR *q* values, respectively, computed on the meta-analysis results of the relevant trait. HeartBEAT replication *p* values are one-sided *p* values guided by the direction of association in the meta-analysis. Fig. 2 visualizes the effect sizes for these transcripts in MESA and FOS for all investigated traits.

**Fig. 2.**
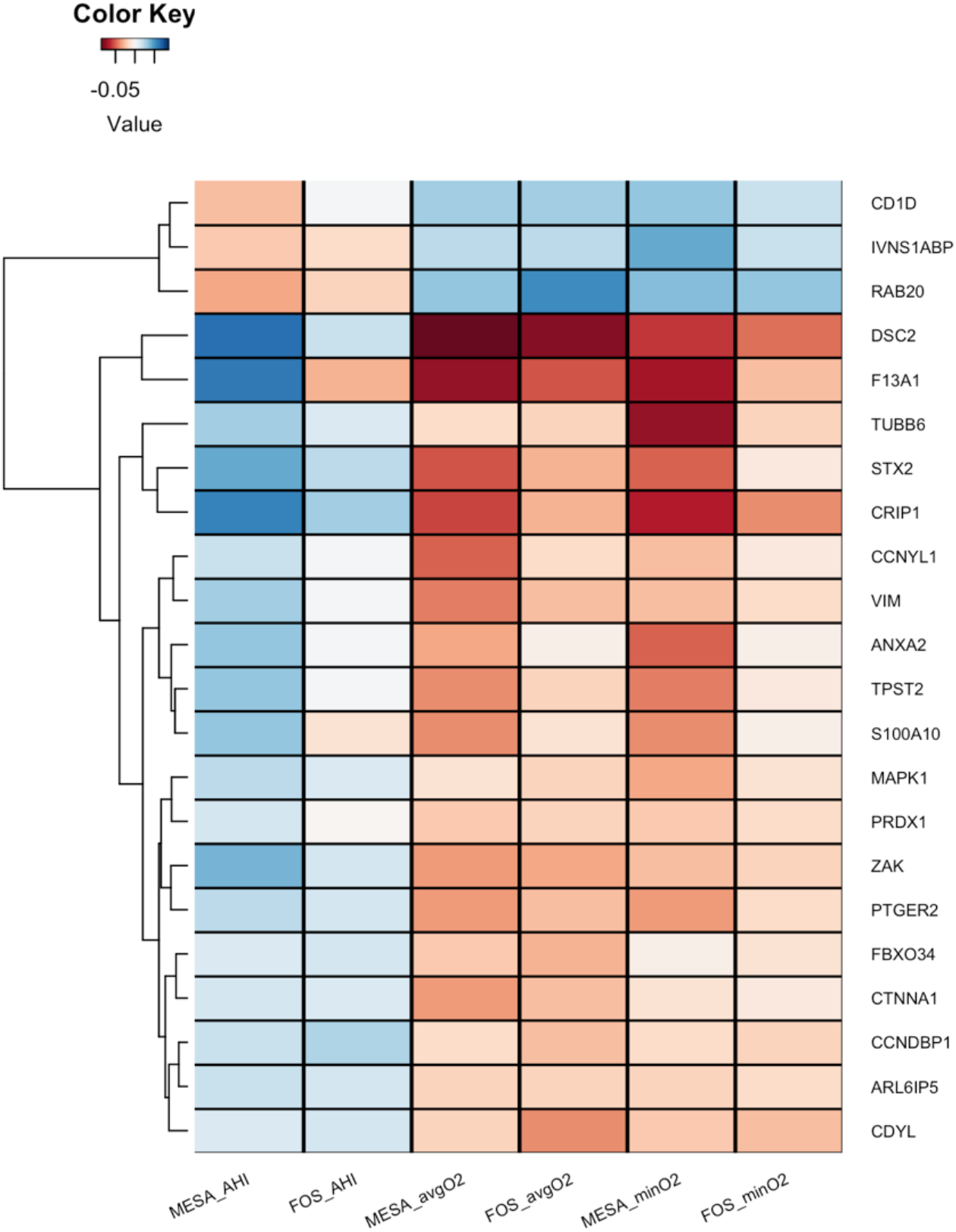
Clustered heatmap of standardized effect sizes for the OSA-associated genes in each of the OSA traits. For each of the genes that cross-replicated in MESA and FOS, or were discovered in their meta-analysis, the figure displays the effect sizes per 1 standard deviation of the trait, in the study. In MESA, effect sizes are based on the meta-analysis of the estimates in the three race/ethnic groups. Positive (blue) effect sizes of AHI and negative (orange) effect sizes of avgO2 and minO2 represent higher expression with more severe OSA symptoms. When multiple probes interrogated a single gene, the figure displays only one of them, selected at random, because the results were similar across all transcripts within each gene.

### Pathway-based identification of processes altered in SDB

To gain a more comprehensive assessment of gene expression patterns of SDB-associated traits, we complemented our gene-focused association analysis by applying Gene Set Enrichment Analysis (GSEA) to the entire available transcriptome in each cohort. We surveyed 1,379 canonical pathways and applied a strict FDR significance threshold (FDR *q <* 0.05). Data file S1 provides complete results and Table S1 provides the number of significant pathways (FDR *q* < 0.05) in each of the analyses. Supplementary Fig. S1 visualizes all pathways with FDR *q <* 0.05 in one analysis and *p* < 0.05 in at least one other analysis.

In the FOS study, in which gene expression was assayed several years after the sleep study, heme metabolism was strongly and significantly up-regulated with respect to SDB severity; this was also observed in MESA, although it did not pass multiple testing correction (AHI *p* = 0.02; avgO2 *p* = 0.002). Other pathways had less consistent patterns across analyses. In MESA, the strongest signal was found for avgO2: 150 gene sets were significantly up-regulated (FDR *q*<0.05) with low average oxyhemoglobin saturation (*i*.*e*., with worse SDB). A visual summary of this result is depicted in Fig. 3, where network analysis was used to link related gene sets into larger modules comprised of functionally similar processes, in particular those related to immunity and inflammation or development and remodeling. In the HeartBEAT cohort, we found that pathways involved in immunity and inflammation, transcription and cell cycle, and circadian rhythms were down-regulated following CPAP therapy, whereas gene sets related to oxidative phosphorylation and metabolism were up-regulated (Fig. 4). Taken together, our pathway analyses across multiple cohorts suggested that immuno-inflammatory processes are activated in individuals with low avgO2 and, importantly, effective treatment of OSA with CPAP moderates this pro-inflammatory signal.

**Fig. 3.**
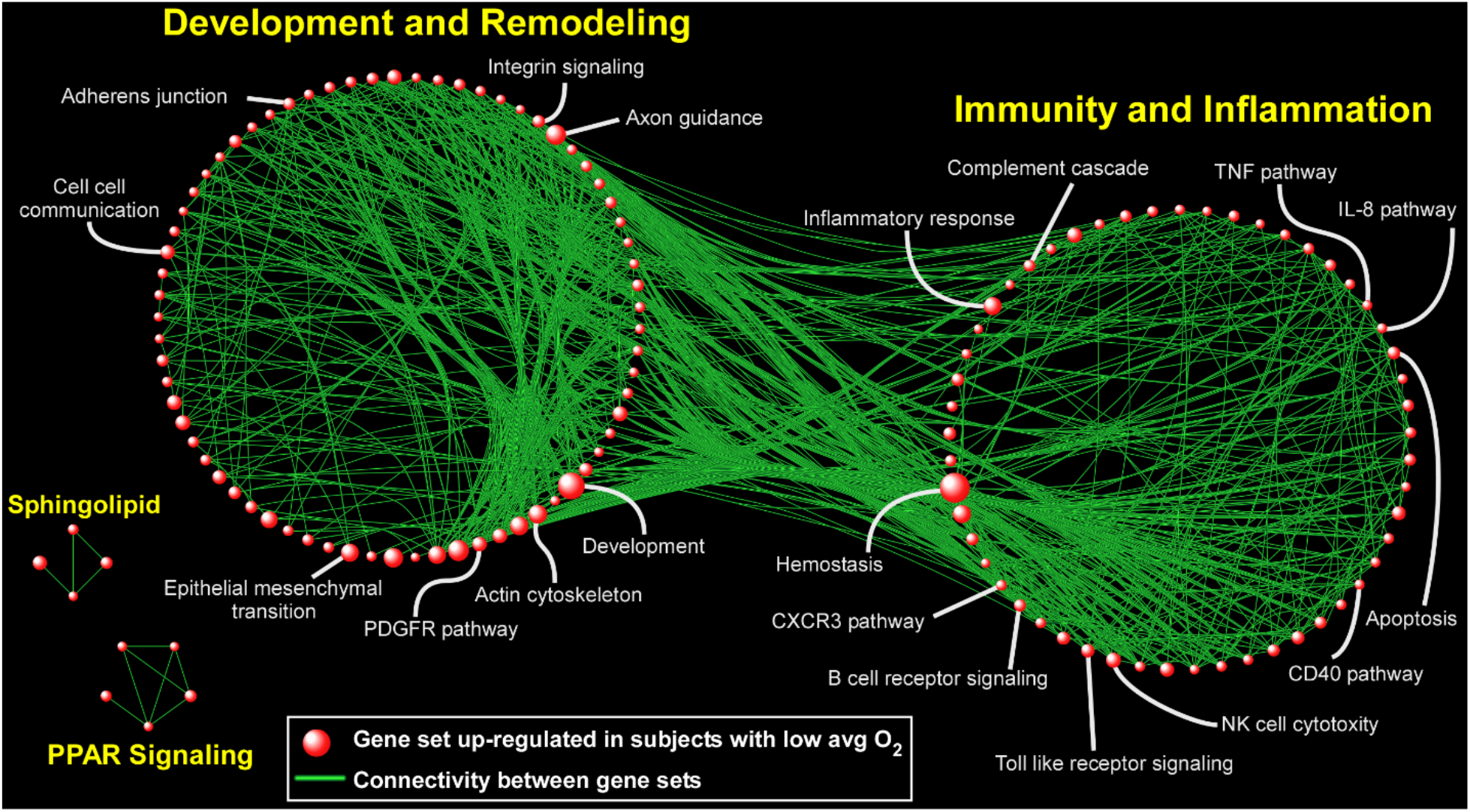
Network-based depiction of up-regulated gene sets in the analysis of gene expression association with lower avgO2 (i.e., more severe SDB) in MESA. Gene sets with FDR *q <* 0.05 are shown. The sizes of the symbols correspond to the number of genes in the set. Connectivity is based on overlapping genes among gene sets. Note that we did not identify any gene sets that were down-regulated (FDR *q* < 0.05) in association with more severe SDB.

**Fig. 4.**
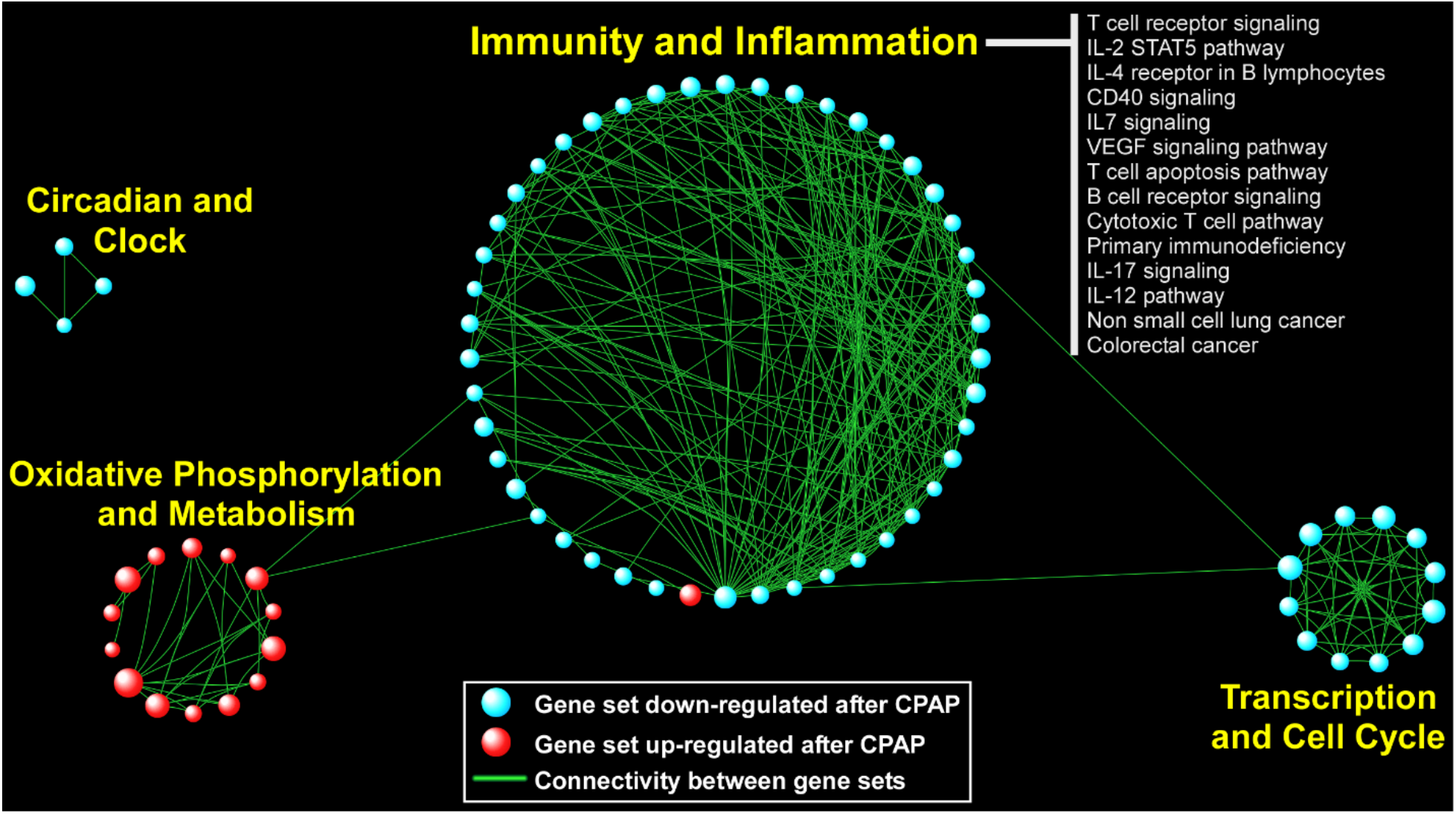
Network visualization of up- and down-regulated gene sets detected in the HeartBEAT study. Gene sets with FDR *q <* 0.05 are shown. The sizes of the symbols correspond to the number of genes in the set. Connectivity is based on overlapping genes between gene sets.

## Discussion

We performed a comprehensive study of the association of SDB traits with genome-wide gene expression in peripheral blood. We combined two large population-based studies, FOS and MESA, to study the associations of three continuous measures of SDB in the general population, and we also compared gene expressions before and after CPAP treatment in a subset of individuals from the HeartBEAT clinical trial who had moderate to severe OSA and were adherent to treatment. We identified 22 genes whose expressions were associated with SDB traits in either cross-replication or meta-analysis of FOS and MESA. Most of these differentially expressed genes were associated with measures of hypoxemia: avgO2 and minO2. While these measures of hypoxemia may be influenced by conditions other than SDB, two of these genes, *CD1D* and *RAB20*, showed evidence of change with OSA treatment in the expected directions; both were expressed at lower levels in individuals with more severe SDB in FOS and MESA, and their expression increased significantly in HeartBEAT patients following treatment with CPAP, suggesting that OSA was the cause of their differential expression.

The *RAB20* gene encodes the Ras-related protein Rab20, a member of the Rab family of small GTPAses. Expression of *RAB20* is regulated by Hypoxia-Inducible transcription factor (HIF-1α) (46), a protein whose expression responds to changes in tissue oxygen tension, and enhances oxygen availability and reduces oxygen demand in a hypoxic microenvironment (47). In our analysis, more severe SDB (higher AHI, lower minO2, and avgO2) was associated with lower expression of *RAB20*. Impaired *RAB20* expression in human skeletal muscle is associated with impaired skeletal muscle glucose uptake and reduced total body insulin sensitivity (47). Reduced insulin sensitivity is a common pathophysiological finding in SDB (48, 49) and can be induced by as little as 8 hours of exposure to intermittent hypoxia (50, 51). Recent animal data also indicate that *RAB2*0 is a negative regulator of neurite outgrowth, and thus may influence neuronal network development, potentially influencing brain development (52). The lower *RAB20* expression in association with SDB severity is somewhat surprising, as hypoxemia is reportedly associated with HIF-1-mediated up-regulation of *RAB20* expression (34,(46). There was only a very weak evidence for association of *HIF-1α* expression with SDB traits in our analysis, however, possibly reflecting the complexity of pathways that regulate HIF-1, including both hypoxia and redox-sensitive pathways, as well as the predominantly post-transcriptional mechanisms regulating HIF-1 signaling (53). That CPAP treatment was associated with increased *RAB20* expression in HeartBEAT subjects implies that OSA was the cause of reduced RAB20 expression in the cross-sectional analysis, and suggests a mechanism whereby CPAP treatment may improve glucose metabolism in OSA patients.

The *CDID* gene encodes the CD1D antigen, an MHC class I-like molecule expressed on both myeloid and lymphoid cells that binds glycolipids and presents them to T cell receptors on natural killer T cells (54, 55). It is possible that reduction in CD1D antigen, potentially associated with the observed reduction in its expression, may contribute to impaired immune function that has been observed in individuals with OSA (56, 57). *CD1D* expression was reported to be regulated by retinoic acid (58) in mononuclear cells. In addition, the retinoic acid pathway was significantly down-regulated in lower avgO2 in MESA, and up-regulated in response to CPAP treatment in HeartBEAT (Data File S1), suggesting a role for retinoic acid in mediating reduced CD1D expression in SDB. It has also been reported that natural killer (NK) T cells have impaired maturation and cytotoxicity in untreated OSA patients, and that this immunosuppressive phenotype was correlated with severity of hypoxemia (59). Given the apparent importance of NK cell activity to immunosurveillance of human cancer (60), CD1D-mediated impairment in natural killer cell function may be implicated in the increased cancer risk in individuals with OSA (61, 62) as well as the increased tumor implantation and growth in animal models of intermittent hypoxia (63, 64).

Other genes identified by the FOS and MESA analysis that did not demonstrate a response to CPAP treatment in HeartBEAT subjects include *MAPK1*, involved in the HIF-1*α* response to hypoxia (65), and *CTNNA1*, that encodes a protein shown to respond to chronic intermittent hypoxia in a mouse study of transcription in cardiac rhythm genetic networks (66). Two related genes were up-regulated under lower oxygen saturation: annexin A2 (*ANXA2*) and S100 Calcium Binding Protein A10 (*S100A10*). These genes code proteins that interact with each other (67) and both were shown to contribute to pulmonary microvascular integrity by preventing vascular leak during alveolar hypoxia in mice (68). Since the expression of these genes did not respond to CPAP therapy, they may indicate persistent alterations in transcriptional response as a result of chronic OSA.

Hypoxemia is known to influence the expression of a wide array of genes and prior studies have demonstrated many more genes are up-regulated than down-regulated in response to experimental sustained or intermittent hypoxemia (50). Consistent with this finding, we observed many more pathways up-regulated than down-regulated in association with increased severity of SDB traits such as low avgO2. Gene set enrichment analysis demonstrated up-regulation of heme metabolic pathways across FOS and MESA cohorts with more severe SDB symptoms. Previous experiments showed that subjecting cells to hypoxia up-regulates the expression of several proteins involved in iron/heme metabolisms, including transferrin (69), transferrin receptor (70), ceruloplasmin (71), and erythroid 5-aminolevulinate synthase (72) in order to increase erythropoiesis and hematopoietic iron supply. Our previous genetic association analysis identified variants eQTL for ferrochelatase (*FECH*), the terminal enzyme in heme biosynthesis, associated with SDB traits (73), suggesting a possible bidirectional association between SDB and heme metabolism pathways.

A key finding of our study is that in a large, ethnically diverse subject cohort to date (MESA), was demonstrating that SDB is associated with up-regulation of pathways involved in immune function and inflammation (Fig. 3). Critically, we found that this immuno-inflammatory signal was down-regulated in the HeartBEAT study patients following CPAP treatment (Fig. 4), providing the first transcriptome-wide evidence that is consistent with a reversal of known pro-inflammatory markers of OSA with effective therapy (74). Another large biological module emerging from pathway analysis of the MESA cohort was up-regulation of gene sets involved in development and remodeling. This finding is supportive of the growing evidence based on animal models and human data that OSA and nocturnal hypoxemia are associated with aberrant cardiovascular remodeling (75-77). In the HeartBEAT cohort, we also observed down-regulation of pathways involved in cancer and cell cycle following resolution of OSA and nocturnal hypoxemia with CPAP therapy. Although many cell types, including lymphocytes, may undergo cell cycle arrest in response to hypoxia (78, 79), our findings are consistent with recent work supporting down-regulation of neoplastic programs after CPAP (23) and the potentially increased risk of cancer associated with OSA and nocturnal hypoxemia (80, 81).

In this work, we utilized data from three different studies to identify genes and gene sets that show expression levels that vary with SDB traits, then explored which expression signals change with OSA treatment. These studies differed in designs and populations. Notably, despite differences in the temporal associations between the sleep studies and gene expression assays (in MESA the sleep study was performed during the year following the blood draw while in FOS, the sleep study was performed between 13 to 2 years prior to the blood draw), and racial/ethnic composition of the samples, we observed consistent associations for 22 genes in FOS and MESA. The comparability of findings despite variations in the timing between the sleep phenotyping and blood draws may reflect that relative stability of SDB traits over approximately 5 years (82). In the Supplementary Materials, we further report look-ups of previously-reported genes associated with OSA and with CPAP treatment.

These analyses in the observational MESA and FOS studies cannot distinguish between effects of OSA on gene expression and effects of gene expression as a cause of OSA. As the associations observed are strongest for measures of sleep-related hypoxemia, rather than the AHI, these analyses are also unable to distinguish effects of OSA from effects of hypoxemia that may be due to underlying pulmonary disease. The data from HeartBEAT study showing that the reduced gene expression of *CD1D* and *RAB20* seen in the cohort studies was reversed by CPAP treatment is strong evidence that OSA is the cause of the reduced expression of these genes. However, the HeartBEAT sample was small and the lack of significant change in gene expression for the other signals that were identified in the FOS and MESA discovery step does not exclude an etiological role of those genes in OSA-related regulatory pathways.

SDB is strongly associated with BMI (83), and it is challenging to disentangle the effect of SDB from that of BMI. In a sensitivity analysis (Tables S2 and S3), most of the associations become slightly weaker after adjustment for BMI; however, BMI directly influences SDB by altering airway collapsibility, and it is therefore not clear that adjustment for BMI results in a less rather than a more confounded effect estimate. The response to CPAP therapy of *CD1D* and *RAB20* indicates that the expression changes identified for these genes is due to OSA and not obesity.

In summary, we report the largest gene expression analysis of SDB traits to date. While precise measures of SDB are challenging to obtain in large-scale cohorts, we leveraged the availability of population-based studies that performed in-home polysomnography and combined our transcriptional analysis of these cohorts with a clinical trial for better control of confounding and confirmation of expression changes with treatment. Using a gene-focused discovery and validation methodology, we identified *CD1D* and *RAB20* as being key down-regulated genes in SDB whose expression responded to CPAP treatment. A complementary pathway-based approach revealed that SDB severity is associated with alteration of distinct peripheral blood programs including up-regulation of immuno-inflammatory signals that were reversed with effective CPAP therapy. Collectively, our study provides a framework to investigate the transcriptional consequences of SDB traits in multi-cohort datasets and identify putative candidate genes and processes driving this response.

## Data Availability

All summary statistics from analysis reported in this manuscript are provided in the Supplementary Material. Raw gene expression data for FOS are a part of SABRe CVD study on dbGaP, with dbGaP study accession: phs000363.v17.p11. Data could could be obtained by requesting authorized access to the Framingham Cohort Study phs000007 on dbGaP. Gene expression data for MESA could be obtained using application to the MESA https://www.mesa-nhlbi.org. Raw and processed gene expression data for HeartBEAT have been deposited in the Gene Expression Omnibus (GEO), Series GSE133601.

## Supplementary Materials include

Supplementary Methods

Fig. S1. Overlap between enriched gene sets at the *p* < 0.05 level.

Table S1. Number of enriched gene sets in each of the cohort and trait analyses.

Table S2. Results from sensitivity analysis in MESA, for transcripts reported in Table 2.

Table S3. Results from sensitivity analysis in MESA, for transcripts reported in Table 3.

Data file S1. Complete results from Gene Set Enrichment Analyses.

Data file S2. Gene expression associations of previously-reported genes.

Data file S3. Complete results from transcriptome-wide association analyses with SDB traits in MESA.

Data file S4. Complete results from transcriptome-wide association analyses with SDB traits in FOS.

Data file S5. Complete results from the meta-analyses of transcriptome-wide association analyses in MESA and FOS.

Data file S6. Complete results from transcriptome-wide association analysis with CPAP treatment in HeartBEAT.

## Acknowledgments

The authors thank the investigators, the staff and the participants of the MESA study for their valuable contributions. A full list of participating MESA investigators and institutions can be found at http://www.mesa-nhlbi.org.

## Author contributions

TS, DJG, and SAG conceptualized the association study. TS, RL, RJ, HL, ACG, NK, BEC, JL, and SW performed statistical analysis and data harmonization. (data collection; study design), all authors critically reviewed the manuscript. YL, JR, and SR collected data and designed components of MESA and its gene expression study. DL collected data and designed components of FOS and the SABRe CVD initiative which collected genes expression data for FOS. RM, SRP, SFQ, SR, and DJG designed and executed the HeartBEAT study, and DJG and AS designed its gene expression study.

## Data and materials availability

all summary statistics from analysis reported in this manuscript are provided in the Supplementary Material. Raw gene expression data for FOS are a part of SABRe CVD study on dbGaP, with dbGaP study accession: phs000363.v17.p11. Data could could be obtained by requesting authorized access to the Framingham Cohort Study phs000007 on dbGaP. Gene expression data for MESA could be obtained using application to the MESA https://www.mesa-nhlbi.org. Raw and processed gene expression data for HeartBEAT have been deposited in the Gene Expression Omnibus (GEO), Series GSE133601.

